# Oral, intestinal, and pancreatic microbiomes are correlated and exhibit co-abundance in patients with pancreatic cancer and other gastrointestinal diseases

**DOI:** 10.1101/2020.01.30.20019752

**Authors:** Mei Chung, Naisi Zhao, Richard Meier, Devin C. Koestler, Erika Del Castillo, Guojun Wu, Bruce J. Paster, Kevin Charpentier, Jacques Izard, Karl T. Kelsey, Dominique S. Michaud

## Abstract

Oral microbiota are believed to play important roles in systemic diseases, including cancer. We collected oral swabs and at least one pancreatic tissue or intestinal samples from 52 subjects, and characterized 16S ribosomal RNA genes using high-throughput DNA sequencing in a total 324 samples. We identified a total of 73 unique Amplicon Sequence Variants (ASVs) that were shared between oral and pancreatic or intestinal samples. Accounting for pairing and within-subject correlation, 7 ASVs showed significant concordance (Kappa statistics) and 5 ASVs exhibited significant or marginally significant Pairwise Stratified Association (PASTA) between oral samples and pancreatic tissue or intestinal samples. Of these, two specific bacterial species (*Gemella morbillorum* and *Fusobacterium nucleatum subsp. vincentii*) showed consistent presence or absence patterns between oral and intestinal or pancreatic samples. Lastly, our microbial co-abundance analyses showed several distinct ASVs clusters and complex correlation-networks between ASV clusters in buccal, saliva, duodenum, jejunum, and pancreatic tumor samples. Toghether, our findings suggest that oral, intestinal, and pancreatic microbiomes are correlated and bacteria of oral origin exhibit co-abundance relationships and demonstrate complex correlation patterns in the intestinal and pancreatic tumor samples. Future prospective studies should aim to uncover the co-abundance of specific microbial communities for studying etiology of microbiota-driven carcinogenesis.

## Introduction

The oral cavity is a major gateway to the human body. It is estimated that the oral cavity collectively harbors over 700 predominant bacterial species ^1^. Oral microbes have been shown to contribute to a number of oral diseases, including tooth caries, periodontitis, endodontic infection, alveolar osteitis, and tonsillitis. It is hypothesized that oral opportunistic or pathogenic bacteria can enter into the blood circulation, passing through the oral mucosal barrier, potentially resulting in abnormal local and systemic immune and metabolic responses ^2^. Oral microbiota are believed to play important roles in systemic diseases such as cardiovascular diseases, diabetes mellitus, respiratory diseases, and cancer ^3-6^.

Many studies have investigated the relationship between the oral or gut microbiome and various cancer risks using different methods and study designs ^7,8^. Among these, colorectal cancer (CRC) is the most studied cancer. The unexpected finding that species of *Fusobacterum*, particularly the oral species *Fusobacterium nucleatum*, are very prevalent (about 30%) in CRC cases suggested an association between the oral microbiota in the colon and CRC ^8^. Research on the relationships between oral bacteria and pancreatic cancer risk stems from a number of observational studies that have reported a higher risk of pancreatic cancer among individuals with periodontitis, when compared to those without periodontitis ^9-11^. A number of studies have examined the association of the oral microbiome with pancreatic cancer risk ^12-15^, but results were inconsistent partially due to differences in methods and study designs. Although one recent study found suggestive evidence that oral dysbiosis is a causative effect of early pancreatic cancer ^12^, more prospective studies are needed to replicate and confirm their findings. Defining oral microbial profiles as non-invasive biomarkers for pancreatic cancer could help screening of high-risk populations.

In an effort to address the specific question of whether the pancreas has its own microbiome, we recruited subjects with planned foregut surgery to obtain pancreatic tissue samples for 16S rRNA gene microbiome analysis. We reported that bacterial taxa known to inhabit the oral cavity, including several putative periodontal pathogens, were common in the pancreas microbiome ^16^. Moreover, bacterial DNA profiles in the pancreas were similar to those in the duodenum tissue of the same subjects, regardless of disease state, suggesting that bacteria may disseminate from the gut into the pancreas. Thereore, the present study explores the oral microbiome in these same patients, making it a critical first step to understanding whether oral microbiota may be used as non-invasive biomarkers for monitoring the process of pancreatic carcinogenesis or disease progression. To our knowledge, no prior study to date has characterized the overall microbiome at multiple sites in the oral cavity and their correlations with the microbiome in pancreatic tissue and intestinal tissue or surfaces.

## Results

### Population and sample characteristics

The present analysis included 52 subjects (**Table 1**). These subjects were between 31 and 86 years old, and contributed a total 324 samples (52 tongue swab, 54 buccal swab, 35 supragingival swab, 48 saliva swab, 22 duodenum tissue, 34 jejunum swab, and 19 bile duct swab samples, as well as 21 pancreatic duct, 6 normal pancreatic tissue and 33 pancreatic tumor samples). Based on the pathology records, ICD10 codes were assigned to each subject: 24 subjects had pancreatic cancer (ICD10 codes C25.0-C25.9), 8 subjects had periampullary adenocarcinoma (ICD10 codes C24.1) and 4 subject had extrahepatic cholangiocarcinoma (C24.0), 11 subjects had other pancreatic conditions (ICD10 codes K86.0-K86.3), and the remaining 5 had other gastrointestinal conditions. Sixty-two percent of these subjects were ever smokers and most received antibiotics days prior to surgery (after providing oral specimens).

**Table 1.** Characteristics of RIH subjects who had at least one oral sample and at least one intestinal or pancreatic sample

| **<u>RIH Subjects (N=52)</u>** |  |
| --- | --- |
| **Characteristic** | **Mean (SD)** |
| Age (years) | 64.1 (12.5) |
| Body mass index (kg/m<sup>2</sup>) | 26.3 (4.9), n=51 |
| **<u>n (%)</u>** |  |
| **<u>Sex</u>** |  |
| Male | 24 (46) |
| Female | 28 (54) |
| **<u>Race</u>** |  |
| Caucasian | 49 (94) |
| Black | 1 (2) |
| Asian | 2 (4) |
| **<u>Smoking status</u>** |  |
| Ever smoker | 32 (61.5) |
| Nonsmoker | 19 (36.5) |
| Missing | 1 (2) |
| **<u>Chemotherapy</u>** |  |
| Never | 37 (71.2) |
| Prior to past 6 months | 5 (9.6) |
| In past 6 months | 6 (11.5) |
| Missing | 4 (7.7) |

SD = standard deviation

A total of 4077 unique Amplicon Sequence Variants (ASVs) were identified across oral swab samples, and 4304 ASVs were identified across pancreatic tissue or intestinal tissue and swab samples via DATA2. ASVs are also referred as “features” in QIIME2 processing, and can be roughly understood as unique identifiers reaching up to the levels of bacterial strains or a group of highly similar strains.

### Microbiome communities at different sites

Based on Shannon index (alpha-diversity measure) and Bray-Curtis PCoA plot (beta-diversity measure), bacterial communities from tongue and saliva samples appear more similar to each other compared to that from buccal and supragingival samples (**Supplemental material 1**). PERMANOVA tests showed that saliva samples were significantly different from tongue (p=0.005) and from supragingival (p=0.0001) but not different from buccal samples (p=0.02) with regards to beta-diversity measures. Furthermore, buccal samples were significantly different from tongue (p=0.006) but not different from supragingival samples (p=0.387), and tongue were significantly different from supragingival samples (p=0.0001).

Bacterial communities in pancreatic tissue and intestinal tissue or surfaces have been previously described ^16^. Briefly, alpha and beta-diversity analyses did not show any visually apparent clustering by sites (i.e., duodenum tissue, jejunum swab, bile duct swab, pancreatic duct, and pancreatic tissue samples). Similarly, the PERMANOVA test results were not statistically significant (no significant differences between sites).

### Shared ASVs and Taxonomy

A total of 73 ASVs were common between oral (any site) and intestinal or pancreatic samples in at least one subject (**Figure 1; Supplemental table 1**). Taxonomic annotations of these shared ASV indicate that *Streptococcus, Veillonella, Prevotella, Fusobacterium, Gemella, Haemophilus*, and *Rothia* are top 7 frequently shared genera (in descending order) between oral and gut or pancreatic samples. The top 7 most frequently shared species include: *Veillonella parvula, Streptococcus parasanguinis clade 411, Fusobacterium nucleatum subsp. vincentii, Gemella morbillorum, Haemophilus parainfluenzae, Prevotella veroralis*, and *Rothia aeria*.

**Figure 1.**
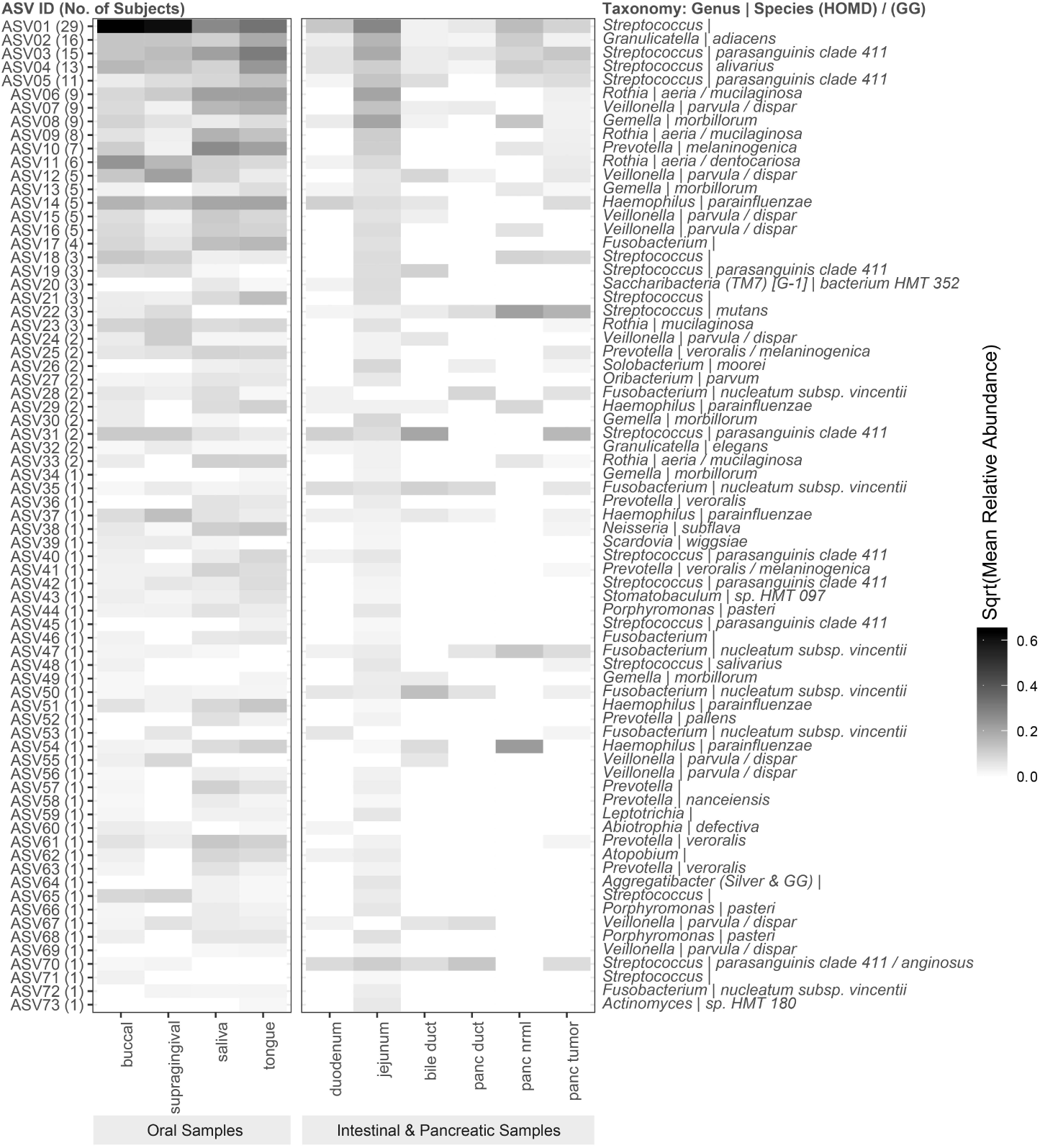
Shared ASVs (number of subjects) and Relative Abundance by Sampling Sites. Legends: On the left panel of the heatmap, each ASV is labeled with a ASV ID and the number of subjects who have the ASV present in both their oral and intestinal & pancreatic samples. Taxonomic annotations of these shared ASV are also provided on the right panel of the heatmap.

### Concordance (Kappa) and Pairwise Stratified Association (PASTA)

For both Kappa and PASTA tests, a total of 53 ASVs for which less than 95% of samples exhibited a relative abundance of zero were tested. Based on the Kappa statistics, seven ASVs showed significant concordance with regards to the probability of presence or absence between oral and pancreatic tissue or intestinal samples after adjusting for multiple testing. The taxonomy of these ASVs (same assigned taxa in all reference databases unless otherwise noted) are: *Oribacterium parvum, Fusobacterium nucleatum subsp. vincentii, Rothia mucilaginosa (GG), Gemella morbillorum, Rothia aeria (HOMD)/mucilaginosa (GG), Streptococcus parasanguinis clade 411 (HOMD)/anginosus (GG), Prevotella veroralis (HOMD)/melaninogenica (GG)*.

However, the estimated probability to be present in both oral and pancreatic tissue or intestinal samples was low (ranging from 2% to 16%) for these seven ASVs (**Table 2**).

**Table 2.**
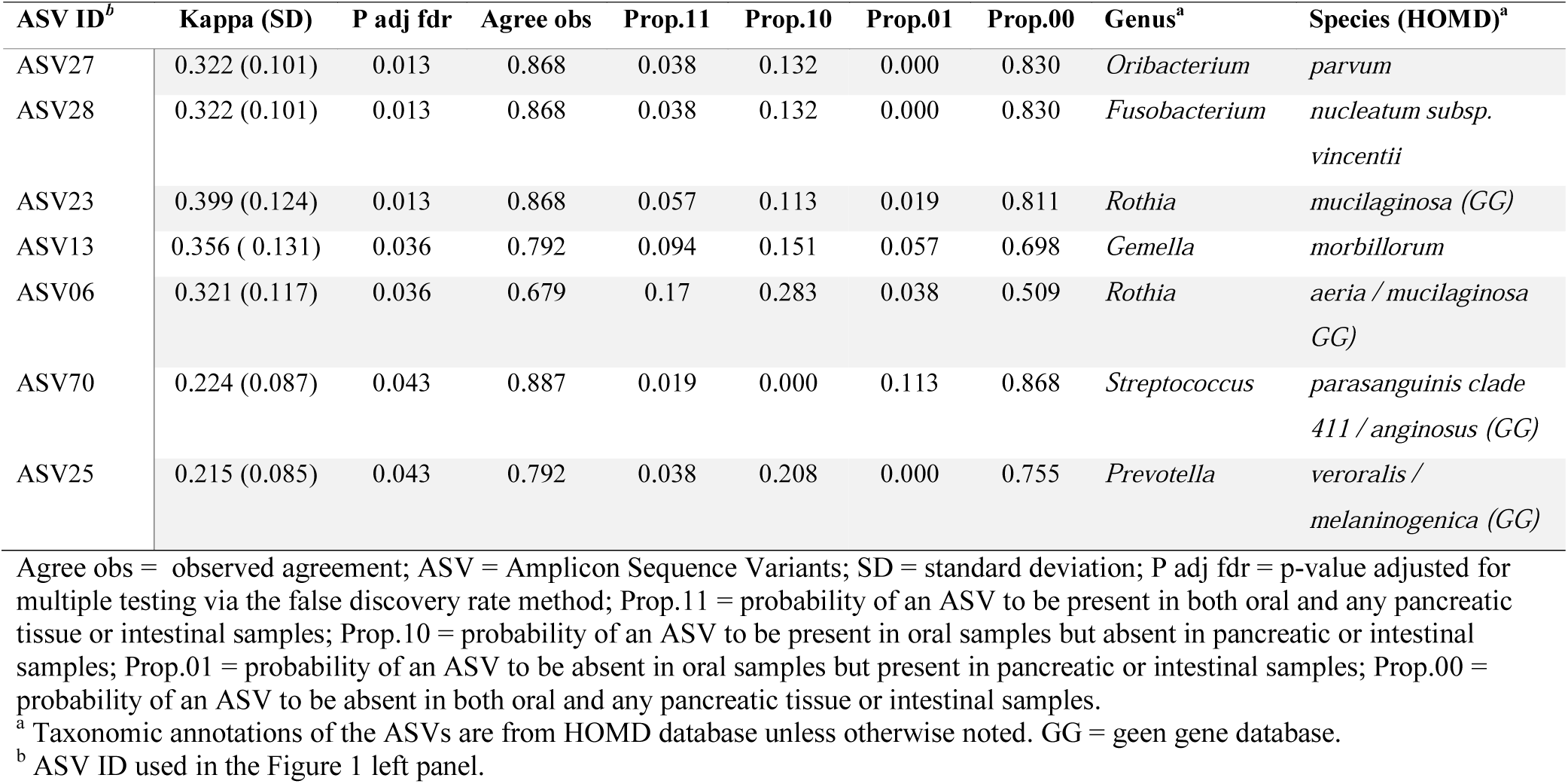
Seven ASVs that showed significant agreement with regards to the probabilities of presence or absence (Kappa statistics) between any oral site and any pancreatic tissue or intestinal samples.

The PASTA test identified two ASVs (ASV13 and ASV21), *Gemella morbillorum and Streptococcus*, that exhibited a significant and a marginally significant association respectively with regards to the probabilities of absence (*p*) between oral and pancreatic tissue or intestinal samples across disease types, when disease status was coded into four groups. For both ASVs, the probabilities of absence tended to be highest among “C24” subjects, second highest among “C25” subjects and lowest among subjects assigned to the “other” or K86.2 group (**Figure 2a**). The ASV corresponding to *Gemella morbillorum* also showed a marginally significant association (PN<0.1) for the mean relative abundance (μ). When disease status was coded into three groups (vs 4 groups), similar findings were shown for the two ASVs in the above analysis. Furthermore, three additional ASVs also showed significant (ASV28: *Fusobacterium nucleatum subsp. vincentii*) or marginally significant (ASV67: *Veillonella parvula/dispar*; ASV19: *Streptococcus parasanguinis clade 411*) associations between oral and pancreatic tissue or intestinal samples with respect to *p* (**Figure 2b**). None of the ASVs tested showed significant associations with regards to the non-zero mean relative abundance (*ω*). Detailed PASTA test results of these five ASVs are shown in **Table 3**.

**Table 3.** Results of the PASTA test when coding disease status into three groups (“C24.*”, “C25.*”, “other”). Five ASVs showed consistent patterns between oral samples and pancreatic or intestinal samples with regards to the probability of absence (*p*), non-zero mean relative abundance (*ω*), or mean relative abundance (μ). The estimated posterior probability of there being no association (PN) between oral samples and pancreatic or intestinal samples is provided in two forms: TP evaluates existence of a linear relationship and TS evaluates the existence of a directional relationship (i.e. consistency of rankings). PN < 0.1 represents moderate evidence of a consistent pattern and PN < 0.05 represents strong evidence of a consistent pattern.

| ASV ID<sup>b</sup> | PN<br>$\mu$<br>TP | PN<br>$\mu$<br>TS | PN<br>$\omega$<br>TP | PN<br>$\omega$<br>TS | PN<br>$p$<br>TP | PN<br>$p$<br>TS | Genus<sup>a</sup> | Species (HOMD)<sup>a</sup> |
| --- | --- | --- | --- | --- | --- | --- | --- | --- |
| ASV28 | NA | NA | NA | NA | 0.0274 | 0.025 | *Fusobacterium* | *nucleatum subsp. vincentii* |
| ASV21 | NA | NA | NA | NA | 0.03 | 0.03 | *Streptococcus* |  |
| ASV13 | 0.0444 | 0.045 | 0.4774 | 0.4744 | 0.0432 | 0.0462 | *Gemella* | *morbillorum* |
| ASV19 | NA | NA | NA | NA | 0.11 | 0.0976 | *Streptococcus* | *parasanguinis clade 411* |
| ASV67 | NA | NA | NA | NA | 0.0952 | 0.102 | *Veillonella* | *parvula* |

NA = not applicable; PN posterior probability of there being no association (PN<0.1 is moderate evidence, PN<0.05 is strong evidence); TP evaluates existence of a linear relationship; TS evaluates the existence of a directional relationship (i.e. consistency of rankings)
<sup>a</sup> Taxonomic annotations of the ASVs are from HOMD database unless otherwise noted.
<sup>b</sup> ASV ID used in the Figure 1 left panel.

**Figure 2.**
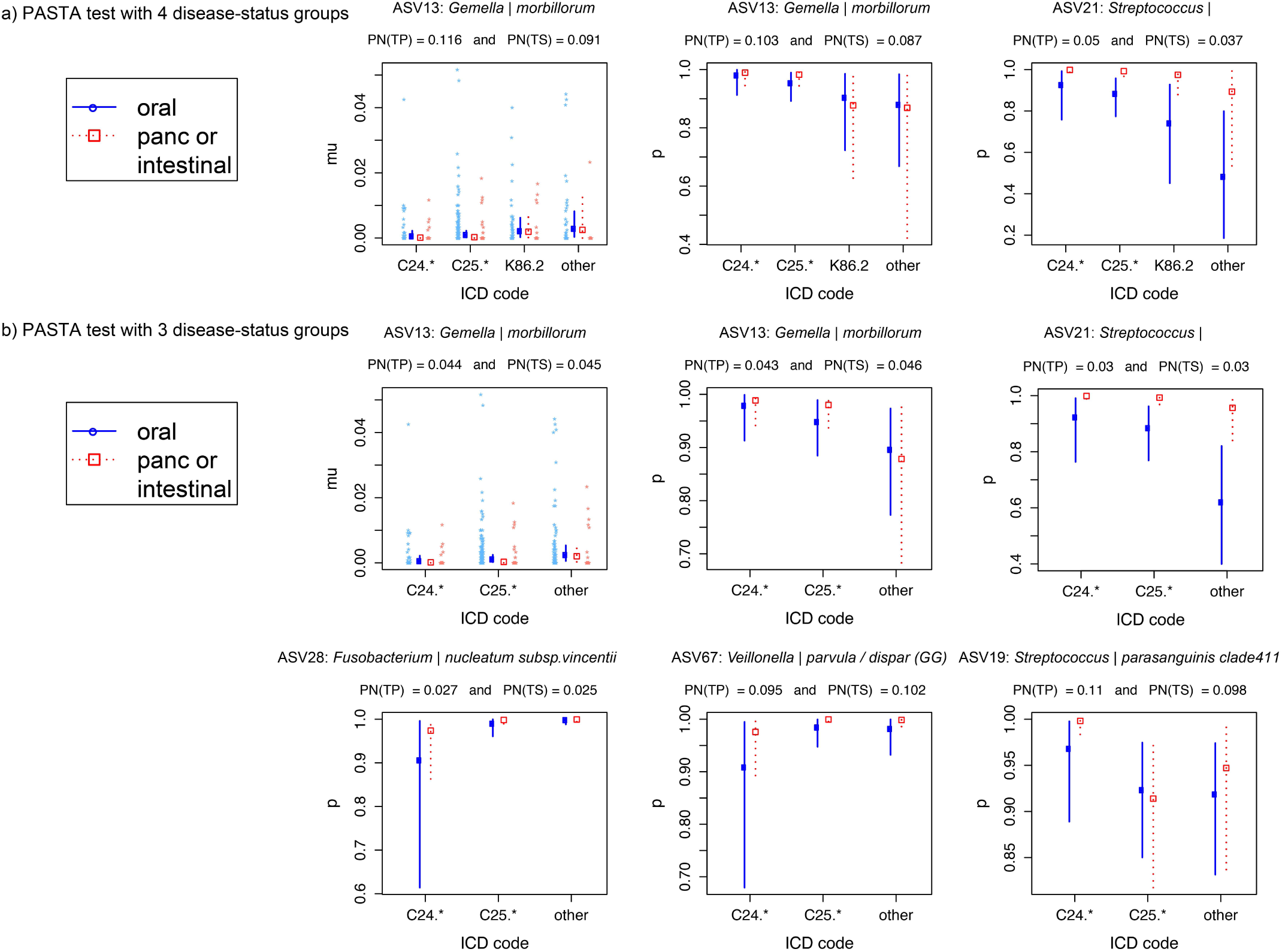
ASVs that exhibited associations between oral samples and pancreatic tissue or intestinal samples after adjusting for disease types and within-subject correlation structure. Disease types were grouped into 4 groups in panel a, and were grouped into 3 groups in panel b. Taxonomic annotations of the ASVs are provided on top of each result plot. Legends: panc or intestinal = pancreatic and intestinal samples; PN = posterior probability of there being no association (PN<0.1 is moderate evicence, PN<0.05 is strong evidence); TP evaluates existence of a linear relationship; TS evaluates the existence of a directional relationship (i.e. consistency of rankings). Plots depict parameter estimates (blue diman or red box) and their 95% credible intervals. For results of the site-specific mean relative abundance (mu), the observed data (blue or red stars) are also plotted next to the intervals.

### Patterns of co-abundance between ASVs

A co-abundance network diagram and clustering tree were graphed for the prevalent ASVs in the saliva (**Figure 3**), buccal (**Figure 4**), duodenum (**Figure 5**), jejunum (**Figure 6**), and pancreatic tumor samples (**Figure 7**), repectively. Only ASVs with an absolute SparCC correlation value greater than 0.1 and a p-value less than 0.05, were plotted. Although ASVs belonging to a co-abundance group does not necessarily imply mutual relationships between them, ASVs in the same co-abundance cluster are likely to respond to environment changes in the same fashion ^17^. In the co-abundance network diagrams, ASVs are colored according to the co-abundance clustering group that they belong to, while their relative positions, ie., being close together or being opposite end of diagram, represent ASVs’ abundance being directly or inversely correlated, respectively.

**Figure 3.**
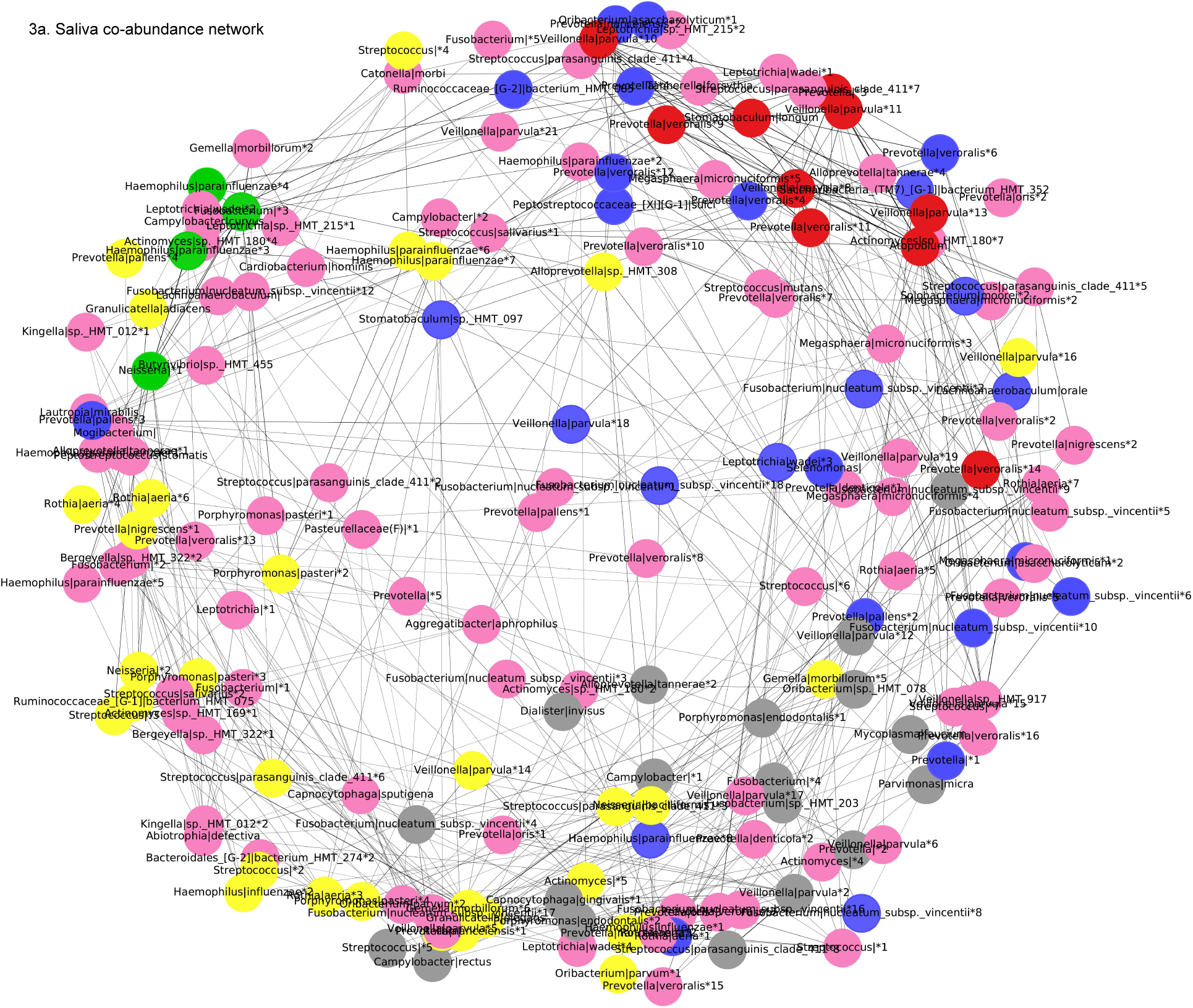
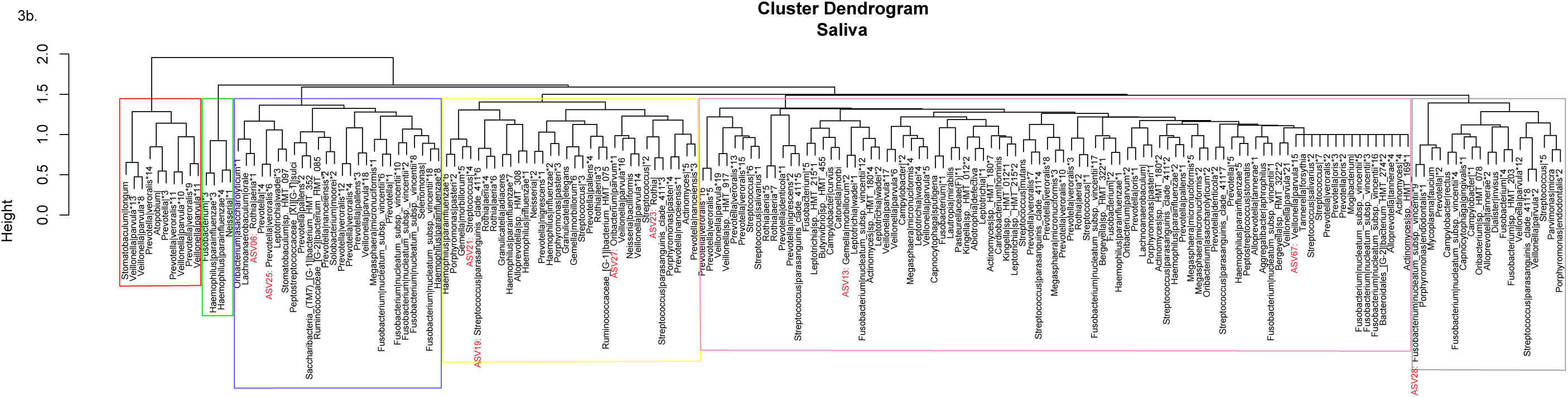
ASVs co-abundance network diagram (panel a) and Ward clusters (panel b) for saliva samples. Legends: Nodes in co-abundance network diagram were colored according to their final co-abundance group assignment described in Ward clusters.

**Figure 4.**
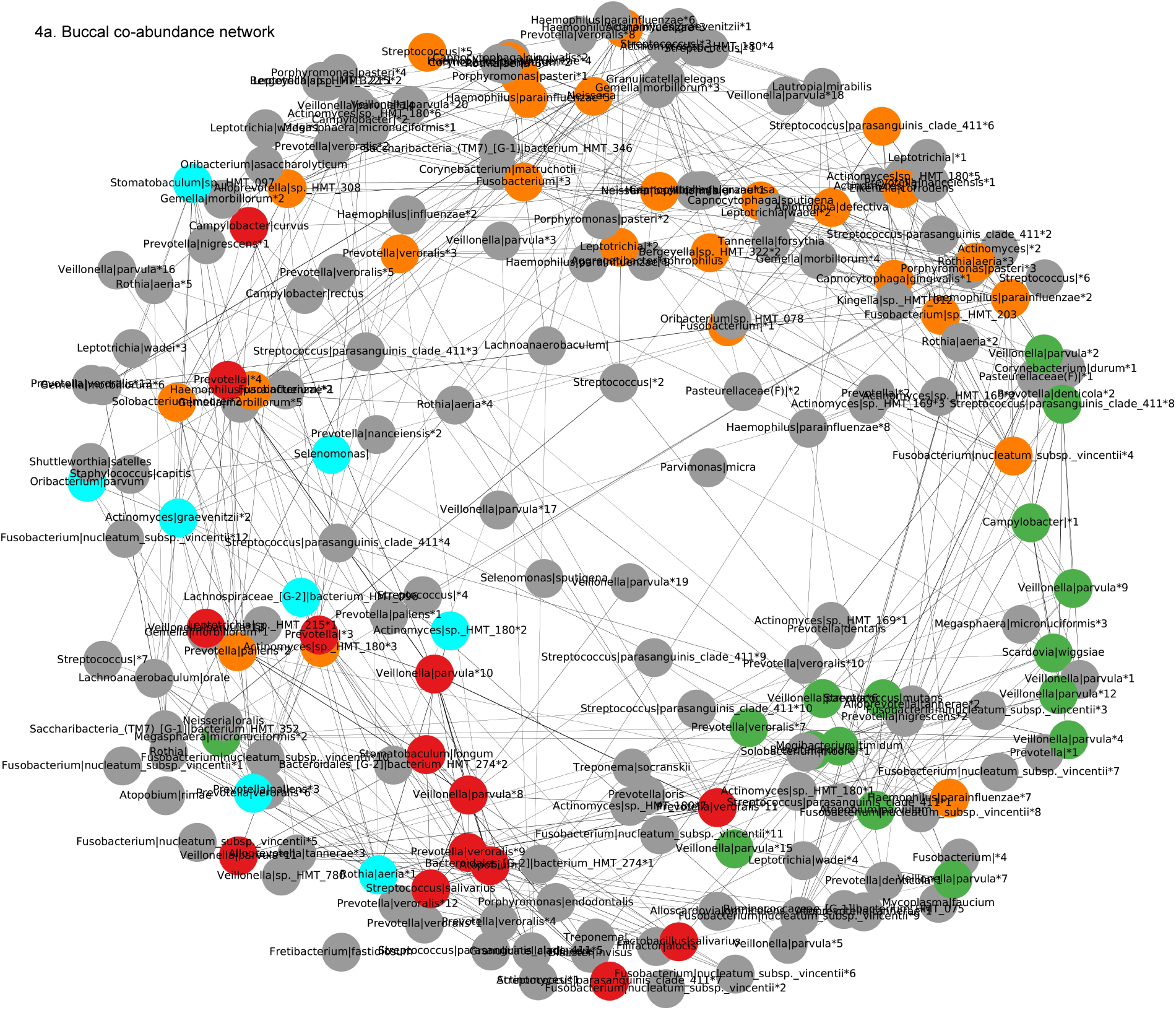
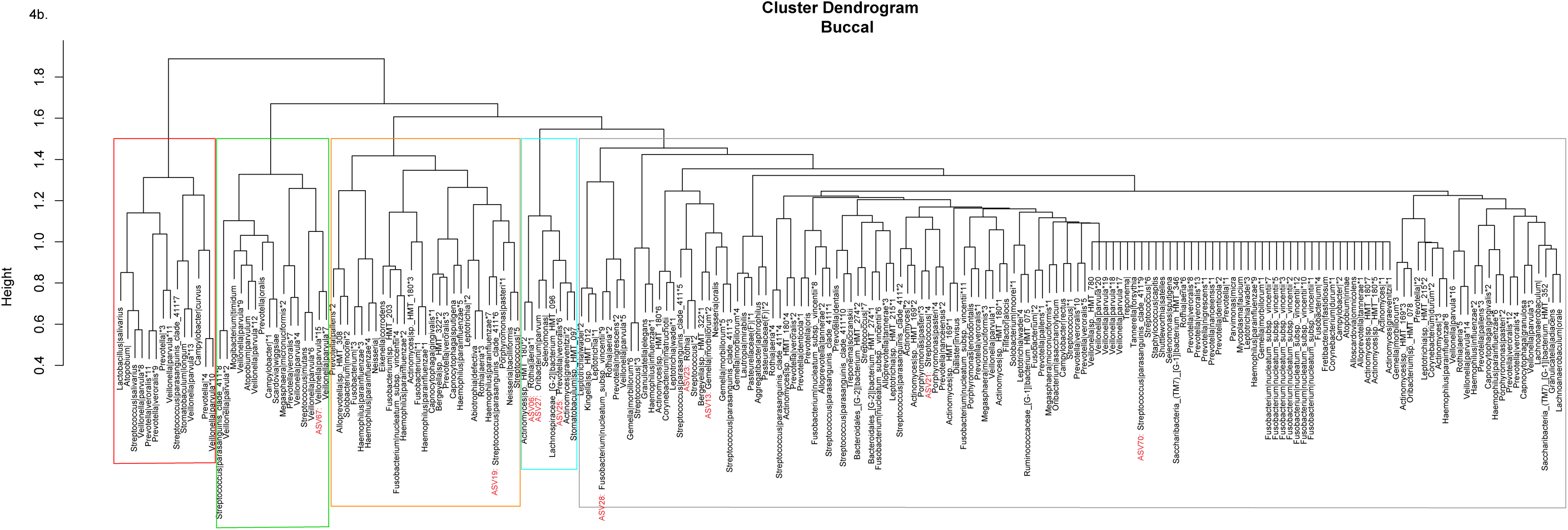
ASVs co-abundance network diagram (panel a) and Ward clusters (panel b) for buccal samples. Legends: Nodes in co-abundance network diagram were colored according to their final co-abundance group assignment described in Ward clusters.

**Figure 5.**
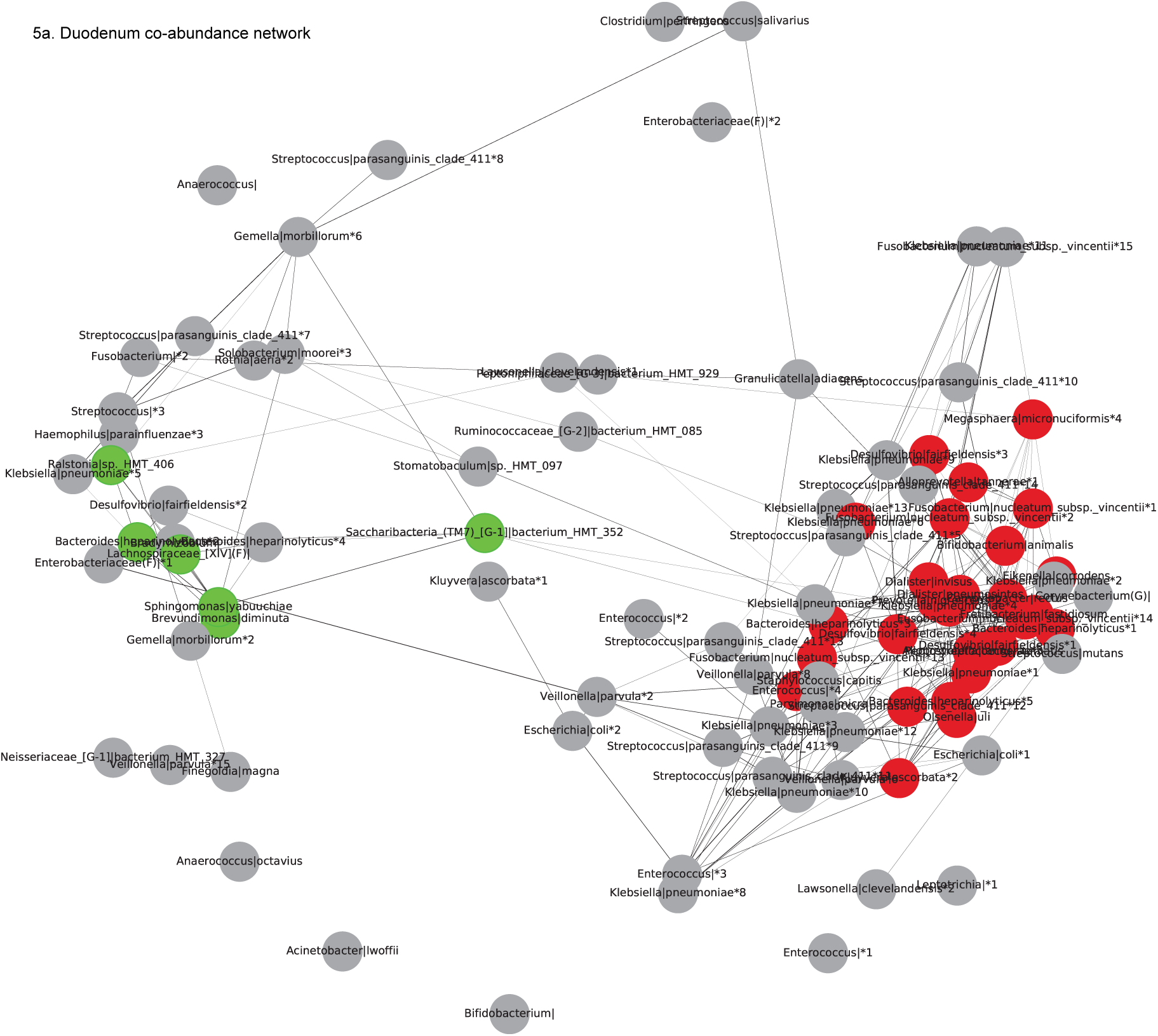
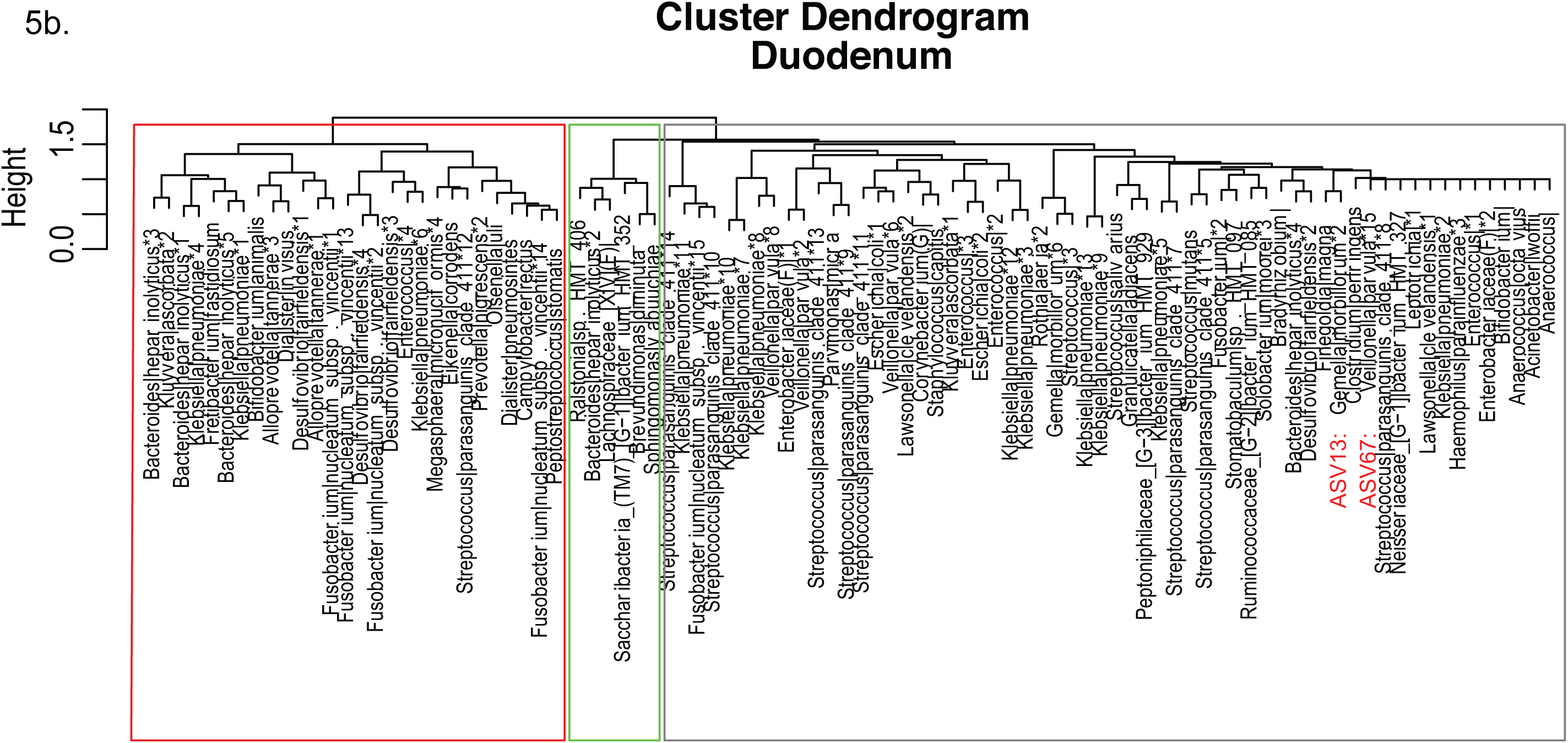
ASVs co-abundance network diagram (panel a) and Ward clusters (panel b) for duodenum samples. Legends: Nodes in co-abundance network diagram were colored according to their final co-abundance group assignment described in Ward clusters.

**Figure 6.**
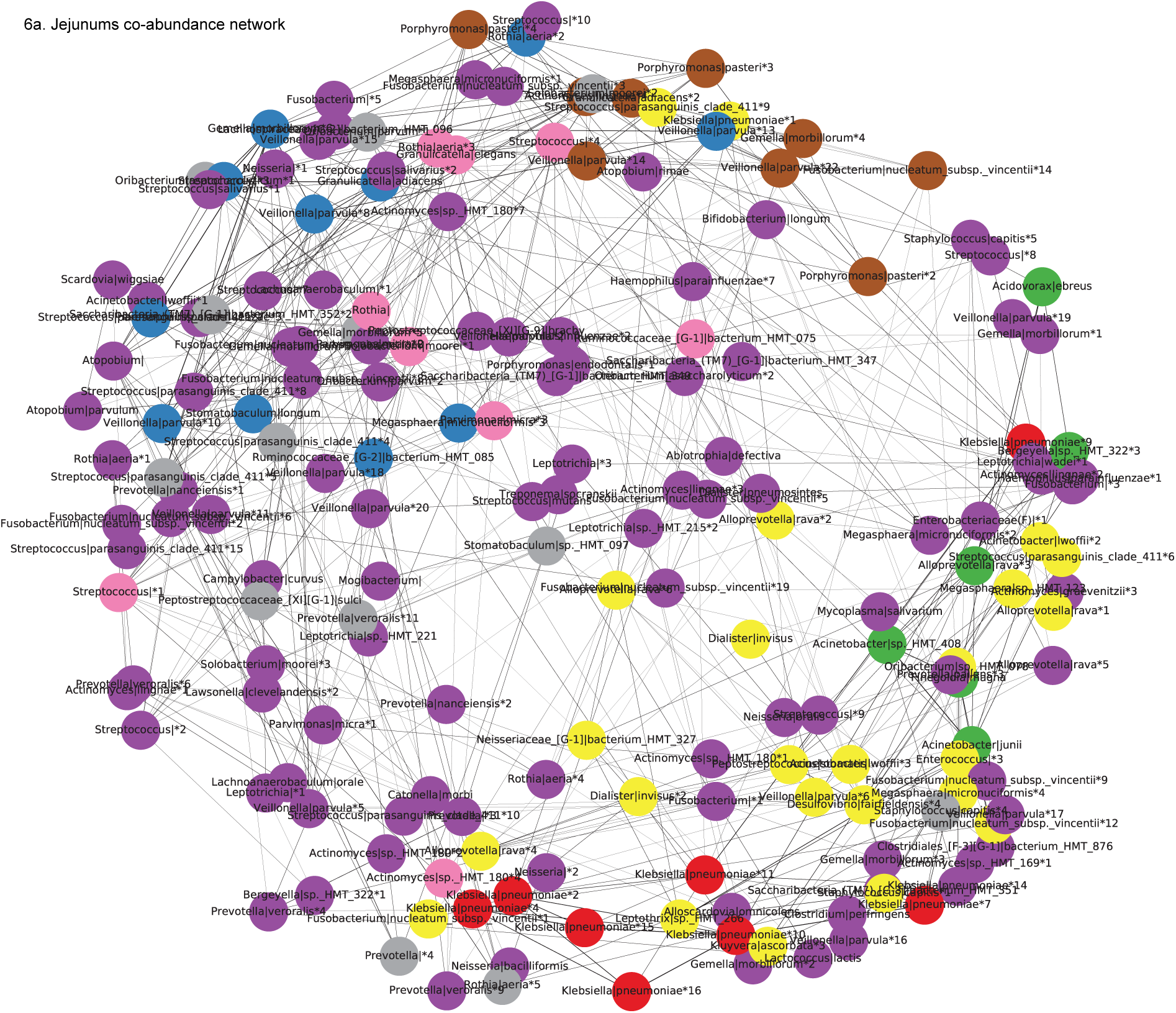
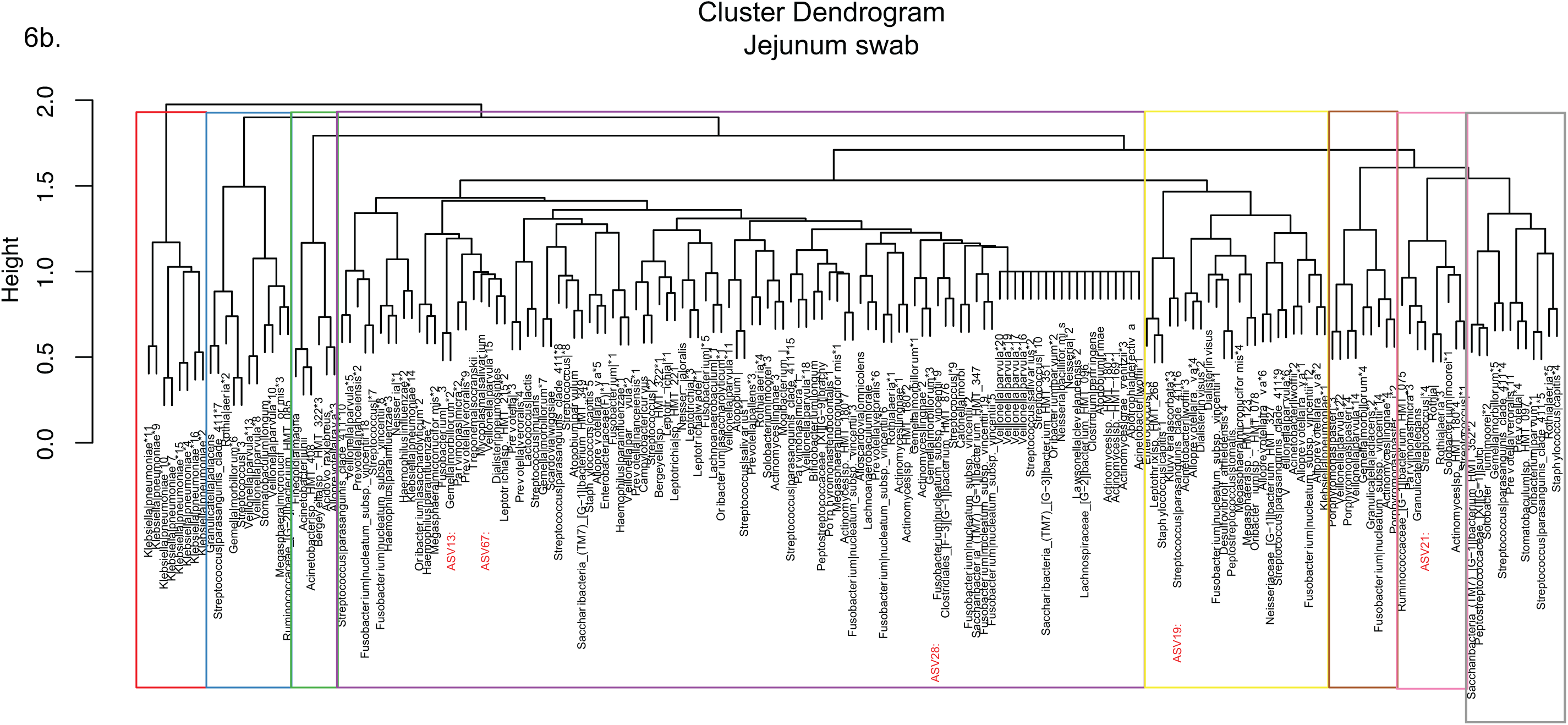
ASVs co-abundance network diagram (panel a) and Ward clusters (panel b) for jejunum swab samples. Legends: Nodes in co-abundance network diagram were colored according to their final co-abundance group assignment described in Ward clusters.

**Figure 7.**
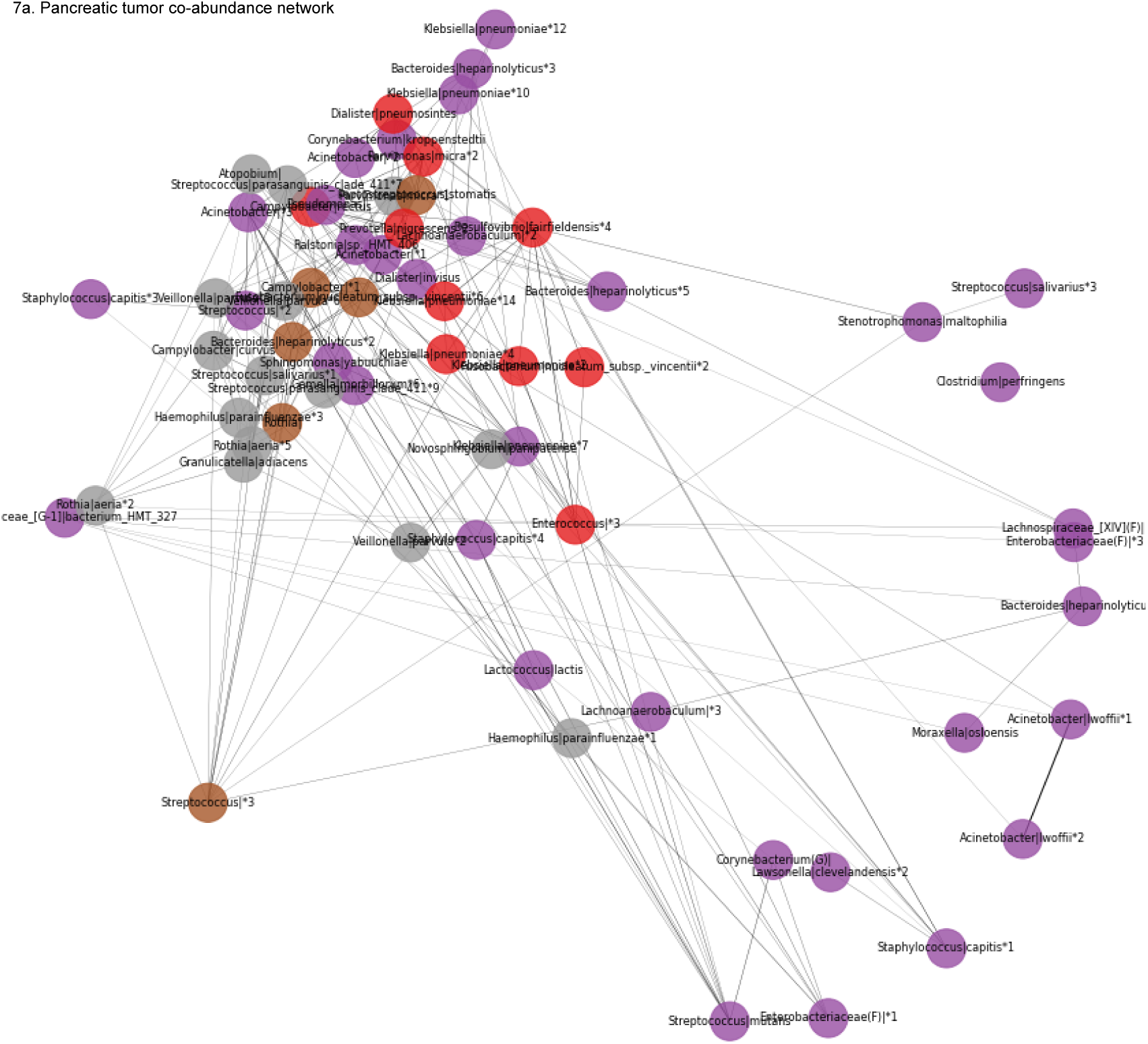
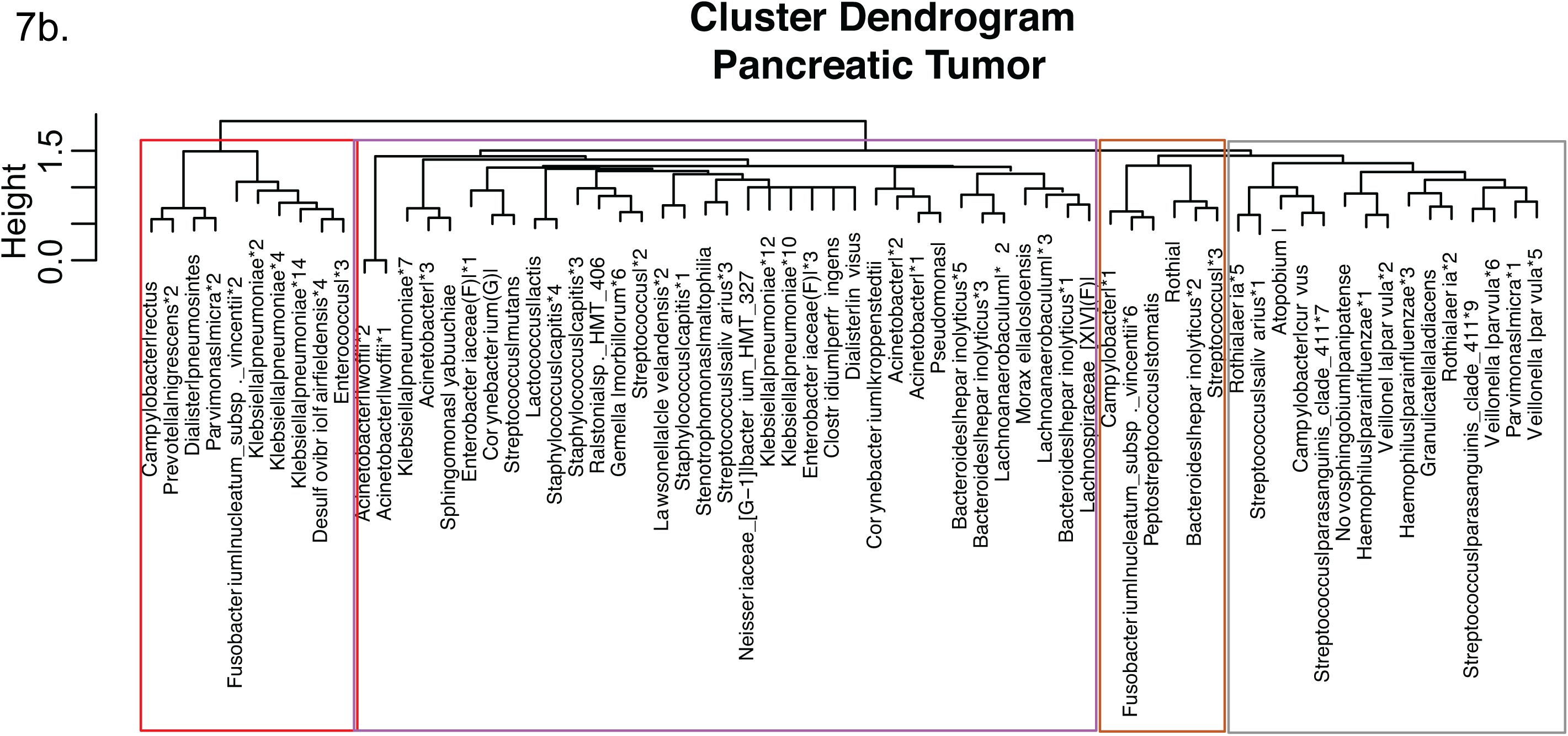
ASVs co-abundance network diagram (panel a) and Ward clusters (panel b) for pancreatic tumor samples. Legends: Nodes in co-abundance network diagram were colored according to their final co-abundance group assignment described in Ward clusters.

The Ward clustering algorithm identified 6 and 5 co-abundance clusters in saliva and buccal samples, respectively (**Figures 3b and 4b**). In buccal samples (**Figure 4b**), the cluster dominated by *Veillonella parvula* (Red and Green) was inversely correlated with the co-abundance cluster that mostly contained *Fusobacterium nucleatum subsp. vincentii, Haemophilus parainfluenzae*, and *Capnocytophaga gingivalis* (Orange). In saliva samples (**Figure 3b**), the same *Capnocytophaga gingivalis* ASV, that was also in buccal samples, was clustered in a co-abundance group with many *Fusobacterium nucleatum subsp. vincentii* and *Streptococcus* ASVs (Grey). Moverover, the co-abundance cluster that contained only *Fusobacterium, Neisseria*|, and *Haemophilus parainfluenzae* (Green) was more positively correlated with a cluster dominated by *Haemophilus* ASVs and *Streptococcus* ASVs (Yellow), and more inverserly correlated with a cluster of *Veillonella parvula* ASVs and *Prevotella veroralis* ASVs (Red). The five ASVs identified by the PASTA tests belonged to the grey cluster (ASV28, ASV21, ASV13, and ASV19) and green cluster (ASV67) in buccal samples. They belonged to the yellow cluster (ASV21 and ASV19), pink cluster (ASV 13 and ASV 67), and grey cluster (ASV28) in saliva samples.

There were 3, 8 and 4 co-abundance clusters in the duodenum tissue, jejunum swab, and pancreatic tumor tissue samples, respectively (**Figures 5b, 6b, and 7b**). All five ASVs identified by the PASTA tests were shown in the jejunum swab clusters – three ASVs (ASV13, ASV28, ASV67) belonged to the purple cluster, one (ASV19) belonged to the yellow cluster, and one (ASV21) belonged to the pink cluster. One duodenum cluster was dominated by *Fusobacterium nucleatum subsp. vincentii* ASVs and *Klebsiella pneumoniae* ASVs (Red), and was inversely correlated to a cluster (Green) that contained *Ralstonia, Bacteroides, Lachnospiraceae, Saccharibacteria, Brevundimonas*, and *Sphingomonas* ASVs. In the pancreatic tumor samples, one cluster was dominated by *Fusobacterium nucleatum subsp. vincentii, Klebsiella pneumoniae, Campylobacter rectus, Dialister pneumosintes, Prevotella nigrescens*, and *Parvimonas* micra ASVs (Red), which contains bacteria that were identified as “the orange complex” – the transitional population between health and severe periodontal disease ^18^. Only two (ASV13 and ASV67) of the five ASVs identified by the PASTA tests were shown in the clusters of ASVs in duodenum samples. Both (*Gemella morbillorum* and *Veillonella parvula*) belonged to the grey cluster. Two ASVs (ASV13: *Gemella morbillorum* and ASV67: *Veillonella*| *arvula*) consistently belonged to the same cluster in saliva, duodenum tissue, and jejunum swab samples. None of the five ASVs were plotted in pancreatic tumor samples’ ASV clusters (due to low or insignificant correlations with other ASVs) although two (ASV13 and ASV28) were present in the pancreatic tumor samples (**Supplemental table 1**).

## Discussions

In this paper, we characterized oral microbiome communities at four oral sites (tongue, buccal, supragingival, and saliva) and examined their correlations with the microbiome in the pancreatic tissue or intestinal samples using several complementary analyses. We identified a large number of ASVs that were shared between oral and pancreatic or intestinal samples among patients with pancreatic cancer and other gastrointestinal diseases. Tongue and saliva samples were more similar to each other with regards to the alpha and beta diversity measures compared to the other two oral sites. This results expand the original finding of the Human Microbiome Project of oral biogeography in healthy subjects ^19^. More importantly, it underlines that individuals with cancer and gastrointestinal diseases maintain such biogeography, offering the option to limit the numbers of oral samples to be collected in future study, in particular longitudinal prospective studies. When comparing oral to intestinal or pancreatic samples, seven ASVs showed significant concordance (Kappa statistics) and five ASVs exhibited significant or marginally significant associations (PASTA) with regards to the probabilities of absence or precence. Lastly, our microbial co-abundance analyses showed several distinct ASVs clusters and complex correlation-networks between ASV clusters in buccal, saliva, duodenum, and pancreatic tumor samples. Analyzing multiple samples within individuals, the present study is the first to show correlations between oral and pancreatic or intestinal microbiome among patients with gastrointestinal diseases or cancer. We believe our study results provide critical insights for future prospective studies to uncover and define the co-abundance of specific oral microbiome communities as non-invasive biomarkers for monitoring the process of pancreatic carcinogenesis or disease progressions.

Emerging research has focused on profiling oral microbiome as potential biomarkers for disease phenotypes in epidemiological studies ^20^ because they are noninvasive, and oral microbiome profiles have shown relative intraindividual stability over time and clear interindividual differences ^21,22^. Although many observational studies have shown that specific oral microbiota in oral cavities and in the fecal samples are associated with oral, head and neck, lung, colorectal, and pancreatic cancer risks ^7,8,23^, data on the associations between oral and tissue microbiome profiles are very limited. To our knowledge, there is only one prior study examined the microbial profiles that were shared between the oral cavity and tissue samples ^24^. Their analyses focused on 17 OTUs that were detected in 37% of both tissue samples (CRC and polyp) and oral swabs, and identified two tumor-associated bacterial co-abundance clusters.

Specifically, one cluster is comprised of oral pathogens previously linked with late colonization of oral biofilms and with human diseases including CRC (e.g., *Fusobacterum nucleatum, Parvimonas micra, Peptostreptococcus stomatis*, and *Dialister pneumosintes*), and the other cluster is comprised of dominant bacteria in early dental biofilm formation including genera *Actinomyces, Haemophilus, Rothia, Streptococcus*, and *Veilonella* ^24^. A letter to the editor reported that *Fusobacteruim nucleatum* was detected in 8 (57.1%) of 14 CRC patients’ tumor and saliva samples, and identical strains were found in 75% of the tumor and saliva samples suggesting that *F. nucleatum* in colorectal tumor originates in the oral cavity ^25^. In the present study, we identified 73 ASVs shared between oral and intestinal or pancreatic (**Figure 1; Supplemental table** 1). Only *F. nucleatum* was identified from the 1^st^ cluster of the work of Flemmer et al. ^24^, however, all except *Actinomyces* was found from the second cluster in our study. We found that *F. nucleatum* was among the top 3 most frequently shared species based on the taxonomic annotations of the shared ASVs between oral and intestinal or pancreatic samples.

Our co-abundance analyses identified multiple clusters of ASVs in buccal, saliva, duodenum, jejunum swab, and pancreatic tumor samples. In buccal and saliva samples, *Capnocytophaga gingivalis* ASV was consistently found in the same co-abundance group with *Fusobacterium nucleatum subsp. vincentii* ASVs. Of the 4 clusters of ASVs in pancreatic tumor samples, one cluster with largest number of ASVs comprised mostly the dominant bacteria in early dental biofilm formation or the genera also associated with relatively healthy tooth pockets ^18^, and the other three clusters each contain bacterial species that are associated with cancer risks such as *Gemella morbillorum* and *Fusobacterium nucleatum subsp. vincentii*, as well as species that are associated with periodontal disease or infections such as *Prevotella nigrescens, Campylobacter rectus, and Klebsiella pneumoniae* ^26,27^.

After adjusting for disease status, our PASTA analyses identified two specific bacterial species (*Gemella morbillorum and Fusobacterium nucleatum subsp. vincentii*) that showed consistent presence or absence patterns between oral and intestinal or pancreatic samples, which warrant further investigations on their associations to the pancreatic cancer risk or progression. A few studies had shown associations between these bacterial species and CRC risk. A large retrospective study found that the risk of CRC was significantly increased in patients with culture-comfirmed bacteremia (presence of bacteria in blood) from *Gemella morbillorum*, previously known as as *Streptococcus morbillorum* (HR = 15.2; 95% CI = 1.54–150), and from *Fusobacterium nucleatum* (HR = 6.89; 95% CI = 1.70–27.9) ^28^. A case-control study in a cohort of individuals undergoing screening colonoscopy found that both *Gemella morbillorum and F. nucleatum* (part of 21 species-level OTUs) were enriched in fecal samples from CRC patients compared to controls ^23^. This study also showed strong co-abundance relationships between *Parvimonas micra, F. nucleatum, and Solobacterium moorei*. Another study analyzed the association of *Fusobacterium* species in pancreatic tumor with patient prognosis and showed significantly higher mortality (poorer prognosis) among pancreatic cancer patients with *Fusobacterium* species-positive tumors than those with *Fusobacterium* species-negative tumors (HR = 2.16; 95% CI = 1.12–3.91) ^29^.

A growing number of studies have investigated the relationship between the oral microbiome and pancreatic cancer risk using different methods and study designs, but the results have been inconsistent and most studies had small sample sizes ^12-15^. The earliest, published in 2012, is a case-control study examining the oral microbiota of individuals diagnosed with either pancreatic cancer or a matched health control ^13^. This study showed an increase in 31 and a decrease in 25 bacterial species/clusters in the saliva of pancreatic cancer patients compared with healthy controls; two oral bacterial candidates (*Neisseria elongata* and *Streptococcus mitis*) were validated in an independent sample population with the finding of significant reductions (P < 0.05; qPCR) in pancreatic cancer subjects compared to controls. A clinic-based case-control study found that pancreatic cancer patients had higher proportions of the genus *Leptotrichia* and lower proportions of *Porphyromonas* and *Neisseria* in saliva, as compared to healthy controls or those with other diseases (P < 0.001). However, this report found that the relative abundances of previously identified bacterial biomarkers (e.g., *Streptococcus mitis and Granulicatella adiacens*) were not significantly different in the saliva of pancreatic cancer patients ^15^. The largest study to date of the oral microbiome and pancreatic cancer capitalized on data collected on patients in 2 large prospective cohort studies to conduct a nested case-control study ^12^. The results showed that presence (vs absence) of *P. gingivalis* and *Aggregatibacter actinomycetemcomitans* in saliva collected prior to cancer diagnosis was associated with 60% and 120% increase in risk of pancreatic cancer, respectively. In contrast, the presence (vs absence) of *Leptotrichia* in saliva was associated with 13% decreased risk of pancreatic cancer ^12^. The most recently published study evaluated the characteristics of the oral microbiota in patients with pancreatic ductal adenocarcinoma (PDAC), intraductal papillary mucinous neoplasms (IPMNs), and healthy controls ^14^. This case-control study found no differences between patients with PDAC and healthy controls, or between patients with PDAC and those with IPMNs, on measures of alpha diversity of the oral microbiota. PDAC patients had higher levels of Firmicutes and several related taxa (*Bacilli, Lactobacillales, Streptococcaceae, Streptococcus, Streptococcus thermophilus*), although, after adjustment for multiple testing, results remained significant at the phylum level only.

Our study has several strengths. We used ASVs from Illumina-scale amplicon data in all analyses without imposing the arbitrary dissimilarity thresholds that define molecular OTUs. ASVs capture all biological variation present in the data, and ASVs are consistently labelled with intrinsic biological meaning as a DNA sequence ^30^. Thus, our study results can be reliably reproduced and validated in future studies. However, our study also had some limitations; our sample size was small and PASTA tests were likely underpowered. Moreover, we did not have samples from healthy controls and could not compare our findings with prior studies due to differences in methods.

## Conclusions

Taken together, the results of the present study suggest that oral, intestinal, and pancreatic microbiomes are correlated, and bacteria of oral origin exhibit co-abundance relationships and demonstrate complex correlation patterns in the intestinal and pancreatic tumor samples. These findings provide critical insights on microbial communities and species that are common in oral cavity, intestinal or pancreatic tissue samples among patients cancer and other gastrointestinal diseases. Though, due to cross-sectional study design, we are unable to make any conclusion regarding their roles in disease progression or whether these bacterial species were colonizing or just passing through various body sites. Our findings should be validated in independent and adequately sized populations with appropriate controls. Moreover, growing evidence have shown that bacterial species may survive, decline, and adapt as interdependent functional groups responding to environmental changes ^17,31,32^. Future studies should aim to uncover the co-abundance of specific microbial communities for studying etiology of microbiota-driven carcinogenesis in prospective and longitudinal studies.

## Methods

### Sampling, DNA extraction and 16S rRNA gene amplicon sequencing

Subjects of the present study were a subset of subjects who underwent surgery for pancreatic diseases or diseases of the foregut, at the Rhode Island Hospital (RIH) between 2014 and 2016. Details have been described in our previous publication ^16^. Briefly, data on participants’ demographics and behavioral factors were collected using a self-administered questionnaire, and pancreatic tissue samples and gastrointestinal swabs were collected during surgery using DNA-free forensic sterile swabs whenever possible to reduce contamination. Oral swabs were collected from participants prior to surgery using sterile cytology brushes which were immediately placed in tubes containing 700 ul RNA later solution after collection. Saliva was collected using saliva kits (OMNIgene OM-501, DNA Genotek). All samples were de-identified and stored at -80°C until processing for DNA extraction. Hypervariable regions of the 16S rRNA gene were sequenced using primers targeting the V3-V4 as paired-end reads on an Illumina platform. Detailes for DNA extraction and sequencing procedures are provided in our previous publication ^16^. Of the 77 enrolled subjects, 52 subjects with both oral swab samples (tongue, buccal mucosa, supragingival, or saliva) and at least one pancreatic tissue (pancreatic duct, normal or tumor pancreas) or intestinal (duodenum tissue, jejunum swab, bile duct swab) sample, were included in the analyses. Samples of the duodenal stent, stomach swab, ileum swab, and pancreatic swab were exluded due to the small number of samples available.

Sequence quality checking and denoising were performed using DADA2 Illumina sequence denoising process ^33^. Human-associated DNA contaminants were screened out using Bowtie2 ^34^. Taxonomic classification, alignment, and phylogenetic tree building were completed using the Quantitative Insights Into Microbial Ecology version 2 (QIIME2) ^35,36^. The sequences of each sample were rarefied to 1200 to even the difference in sequencing depth across both oral and intestinal samples for further analysis. The choice of 1200 as sampling depth was guided by reviewing an alpha rarefaction curve that tested various depths ranging between 500 and 5,000 (**Supplementary Material 2**).

### Assigning taxonomic annotation

To predict the taxonomic groups that are present in each sample, QIIME2 plugin (q2-feature-classifier) was used to train naïve Bayes classifiers using multiple databases as different set of reference sequences. These were the Human Oral Microbiome Database (HOMD) (version 15.1), the Silva (release 132), and the Greengenes (13_8 revision) 99% OTUs (Operational taxanomic units) 16S rRNA gene databases, all trimmed to contain the V3-V4 hypervariable region. HOMD identification was chosen as the default taxonomy. Whenever HOMD, Silva, and Greengenes yielded different taxonomic results at the species level, taxonomic information from all datasets were kept and reported.

### Statistical analyses

Analyses were carried out in QIIME2 (https://qiime2.org), in R ^37^, and in Python. All analyses were conducted using ASV as the unit of observation in order to accurately capture bacterial strain level variations.

#### Alpha & Beta Diversity

For the calculation of alpha and beta diversity measures of oral microbiome, a phylogenetic tree was first created in order to generate phylogenetic diversity measures such as Faith’s Phylogenetic Diversity, unweighted and weighted UniFrac distances ^38,39^. Creating a phylogenetic tree requires multiple sequence alignment, masking, tree building, and rooting. The masking step removes alignment positions that do not contain enough conservation to provide meaningful information (default 40%). Next, evenness and diversity of oral microbiota in each sample was assessed to examine the variation in the microbial profile across different oral sampling sites. Pielou’s Evenness test and Faith’s Phylogenetic Diversity test were calculated using QIIME2 diversity analyses (q2□ diversity) plugin ^40^. Computed distances were then used to generate principal coordinate analysis (PCoA) plots to visualize the arrangement of the samples in the ordination space. PERMANOVA tests ^41^ were conducted to compare beta-diversity measures between sampling sites. Bonferroni correction was used to adjusted for multiple testing, and thus a p-value less than 0.008 was considered significant differene in beta-diversity measures between oral sites.

#### Identifying Shared ASVs

Descriptive analyses were performed to identify shared ASVs between sites. Rarefied features with a relative abundance less than or equal to 0.01 (≤1%) were set to zero. Shared ASVs between any one of the oral sites (tongue, buccal mucosa, supragingival, or saliva) and any pancreatic tissue or intestinal sites (duodenum tissue, jejunum swab, bile duct swab) for each subject were identified. A heatmap of the ASVs that were shared by one or more subjects was generated in R using the packages *intersect* and *ggplot2*.

#### Concordance and Pairwise Stratified Association

Associations of ASVs between oral and pancreatic tissue or intestinal site samples were investigated using two different types of statistical tests that account for pairing and within-subject correlation. For both tests, rarefied features with a relative abundance less than or equal to 0.01 (≤1%) were set to zero (i.e., set to absence), and only ASVs for which less than 95% of samples exhibited a relative abundance of zero were tested.

The first test (Kappa) evaluated general concordance in presence or absence of a given ASV in each subject. Specifically, for each subject, samples were divided into two groups: 1) oral samples, and 2) pancreatic tissue or intestinal site samples. The target ASV was denoted as present in a group if at least one of the samples had a relative abundance value larger than zero, or denoted as absent if all relative abundance values in the group were zero. This reformatted data thus resulted in two values for each subject: 1) whether or not the target ASV was present across oral samples and 2) whether or not the target ASV was present across pancreatic tissue or intestinal sites for that subject. Based on these values, Cohen’s Kappa concordance statistic was computed ^42^ utilizing the R package “irr”, and a one-sided test based on Kappa’s large sample standard deviation was conducted to examine whether there is significant agreement in presence or absence between oral and pancreatic tissue or intestinal site samples for a given ASV.

Additionally, a test for Pairwise Stratified Association (PASTA) was performed to identify those ASVs which exhibit consistent patterns in relative abundance between oral and pancreatic tissue or intestinal site samples after adjusting for disease status (represented by ICD10 codes). PASTA test was based on a Bayesian regression model to obtain Markov-Chain Monte Carlo estimates of abundance among strata, to calculate a correlation statistic, and to conduct a formal test based on its posterior distribution. For the present analyses, two disease-status stratifications were considered: Grouping samples by four ICD10 codes “C24.*”, “C25.*”, “K86.2” and “other” (encodes other gastrointestinal conditions), and grouping samples by three ICD10 codes “C24.*”, “C25.*” and “other” (includes K86.2 and other gastrointestinal conditions). An ASV was defined to exhibit a consistent PASTA pattern, if either one of three possible quantities was associated between oral and pancreatic tissue or intestinal site samples: 1) The probability of absence (*p*), which denotes the probability of observing a relative abundance of zero, 2) The non-zero mean relative abundance (*ω*), which denotes mean relative abundance among those samples in which ASV is present; and 3) The mean relative abundance across all samples (μ).

To conduct the PASTA test, *p, ω*, and μ were first estimated in each site-comparison-by-disease-status subgroup based on a Bayesian Zero-inflated Beta regression model, which was fit utilizing the OpenBUGS statistical analysis software and the R package “R2OpenBUGS” ^43^.

This regression model accounted for within-subject correlation due to repeated measurements via the inclusion of a subject-specific random intercept term. Using the subgroup estimates of *p, ω*, and μ, the posterior probability of exhibiting no association (PN) between the two site-comparison groups was calculated. This probability can be understood as a rejection threshold, where a value of less than or equal to 0.05 was considered statistically significant (i.e., strong evidence of association) and a value of less than 0.1 was considered as marginally significant (i.e., moderate evidence of association). Further information about the data model, approach, and performance of the PASTA test have been published in detail ^44^.

#### ASV co-abundance groups

ASVs shared by more than 10% of the buccal, saliva, duodenum, and pancreatic tumor samples were considered prevalent ASVs. Correlations between these prevalent ASVs within each sampling site were calculated using the SparCC method (Sparse Correlations for Compositional data) ^45^. The statistical significance of these correlation coefficients was assessed using a bootstrap procedure and then converted into a correlation distance matrix. Next, Ward clustering algorithm, a top-down clustering approach, was used to cluster ASVs within each sampling site into co-abundance groups. Starting from top of the Ward clustering tree, Permutational MANOVA (9999 permutations, p<0.001) was used to sequentially test whether any two branches of the tree were significantly different ^46^. ASVs within the same co-abundance group increased or decreased in abundance together.

#### ASV co-abundance network

Bacterial co-abundance networks illustrate how groups of ASVs occupy different niches of the microbial ecosystem. The co-abundance networks were visualized as force-directed network plots using Python package NetworkX (version 2.2) with the spring layout of the Fruchterman-Reingold algorithm (k=0.15) ^47^ for buccal, saliva, duodenum, and pancreatic tumor samples. Only ASVs with an absolute SparCC correlation value greater than 0.1 and a p-value less than 0.05, were plotted. Force-directed algorithms simulate correlation coefficient values as basic physical properties ^48^. Positive and negative correlations are modeled as gravity and repulsion, respectively. ASVs with strong positive correlations between each other are plotted near each other, while negatively correlated ASVs are pushed away from each other. In the co-occurrence network plots, each node represented a unique bacterial ASV. Lines between nodes represented correlations between the nodes they connect; with line width indicated the correlation magnitude. Nodes were colored according to their final co-abundance group assignment, calculated using Ward clustering algorithm described earlier. During graphing simulation, the forces were applied to the nodes, pulling them together or pushing them further apart. This iterative process continued until the system reached equilibrium state (lowest total energy) and the relative position of nodes stopped changing from one iteration to the next.

## Data Availability

Sequence data have been deposited in Sequence Read Archive (SRA)

https://www.ncbi.nlm.nih.gov/sra/PRJNA558364

## Acknowledgements

We thank the participants who graciously enrolled in this study. We thank Ms. Priyanka Joshi for her tremendous help with recruitment of subjects at the RIH, and Dr. Ross Taliano for assisting with the preparation of the tissue specimens in the Pathology Department at the RIH.

## Authors’ contributions

J.I. and D.S.M. designed this study and obtained funding. N.Z., R.M., and D.C.K. developed the methodology used in the analyses. Sample collection and processing were performed by E.C., B.J.P, K.T.K, J.I, D.S.M., and K.C. Analyses and interpretation of data (e.g., statistical analysis, biostatistics, computational analysis were performed by M.C, N.Z., R.M., D.C.K. G.W., and D.S.M. All authors contributed to draft the manuscript and revised it critically.

## Additioanl Information

### Ethics approval and consent to participate

The study was approved by Lifespan’s Research Protection Office for recruitment at RIH, as well as the Institutional Review Boards for Human Subjects Research at Brown University, Tufts University and the Forsyth Institute. An informed consent was obtained from all subjects. All methods carried out were in in accordance with Helsinki Declaration as revised in 2013.

### Consent for publication

Not applicable.

### Availability of data and material

Sequence data have been deposited in Sequence Read Archive (SRA), accessible with the following link after January 1, 2020: https://www.ncbi.nlm.nih.gov/sra/PRJNA558364

### Funding

The research reported in this publication was supported by the NIH/NCI grants R01 CA166150 and P30 CA168524.

